# Fast raster-scan optoacoustic mesoscopy enables assessment of human melanoma microvasculature *in vivo*

**DOI:** 10.1101/2021.05.25.21257525

**Authors:** Hailong He, Christine Schönmann, Mathias Schwarz, Benedikt Hindelang, Andrei Bereznhoi, Annette Steimle-Grauer, Ulf Darsow, Juan Aguirre, Vasilis Ntziachristos

## Abstract

The development and progression of melanoma tumors is associated with angiogenesis, manifesting as changes in vessel density, morphology, and architecture that may extend through the entire skin depth. Three-dimensional imaging of vascular characteristics in skin lesions could allow diagnostic insights not available to the conventional visual inspection. Raster-scan optoacoustic mesoscopy (RSOM) has emerged as a unique modality to image microvasculature through the entire skin depth with resolutions of tens of micrometers, offering new possibilities to assess angiogenetic processes. However, current RSOM implementations are slow, exacerbating motion artifacts and reducing image quality, particularly when imaging melanoma lesions that often appear on the upper torso where breathing motion is strongest. To visualize for the first time melanoma vasculature in humans, in high-resolution, we accelerated RSOM scanning using an illumination scheme that is co-axial with a high-sensitivity ultrasound detector path, yielding 15 second single-breath-hold scans that minimize motion artifacts. Applied to 10 melanomas and 10 benign nevi in humans, we demonstrate visualization of microvasculature with performance never before shown *in vivo*. We show marked differences between malignant and benign lesions, supporting the possibility to use vasculature as a biomarker for lesion characterization. The study points to promising clinical potential for Fast-RSOM (FRSOM) as a three dimensional visualization method that can enable the complete assessment of microvascular parameters of melanoma and improve diagnostics.

## Introduction

Cutaneous melanoma is one of most aggressive and fatal forms of skin malignancies, responsible for over 10,000 deaths annually in the United States alone [1-3]. Early diagnosis followed by rapid and complete surgical excision are essential for improving prognosis [2, 4, 5]. Currently, the diagnosis of melanoma is largely based on clinical assessment using dermoscopy and histological analysis of an excised lesion or biopsy [6-8]. Dermoscopy only provides a two-dimensional superficial assessment of a lesion, which does not extend beyond the papillary dermis due to photon diffusion [1,2]. Furthermore, visual examination is subjective and accuracy depends largely on the experience of the dermatologist [9]. These factors lead to many false-positives and unnecessary biopsies, which are invasive, slow, and costly [5, 10-12]. For example, it has been reported that the number of benign pigmented lesions excised to detect one melanoma varies from 6.3 to 8.7 for dermatologists and from 20 to 30 for general practitioners [13-16].

Malignant melanoma alters the local skin microvasculature in a manner different from benign lesions, with malignant lesions exhibiting higher vessel density [17-20]. As such, non-invasive imaging of morphological features of lesion microvasculature may complement superficial visual inspection for melanoma detection [19-22]. Furthermore, histological studies have reported that vascular changes induced by malignant melanoma can extend through the entire depth of the skin. It was found that tumor vascularity increases significantly with mean tumor thickness from 1 mm to 4 mm [18, 20, 22, 23], while vessel patterns change such that thin melanomas have more homogeneous vasculature and thick melanomas exhibit more chaotic and heterogeneous vasculature [17]. However, current non-invasive imaging methods cannot fully resolve microvasculature throughout the entire depth of the skin, which hinders the full exploitation of tumor vascular features as biomarkers for melanoma detection.

For example, Doppler ultrasound can detect neovascularization of skin tumor lesions with reportedly high specificity for malignancy (90%–100%) but variable sensitivity (34%–100%) [24]. However, Doppler ultrasound only resolves vessels with diameters > 100 µm in lesions with a thickness of more than 2 mm without applying contrast agents (microbubbles), resulting in poor sensitivity for early stage melanoma detection [24, 25]. Dynamic optical coherence tomography (D-OCT) based on speckle variance allows *in vivo* evaluation of skin vascular patterns and has been applied to distinguish melanoma from benign lesions by examining the lesions’ vasculature [17, 26]. However, D-OCT can only reliably visualize vascular patterns of thin lesions up to a maximum depth of about 500 µm mainly due to scattering [17, 27]. This limited depth penetration misses changes in the deep dermal vasculature for melanoma detection.

In contrast, raster-scan optoacoustic mesoscopy (RSOM) images microvasculature at resolutions in the tens of microns through the whole skin depth by combining optical excitation with ultra-broad bandwidth ultrasound transducers [28-32]. Several studies highlight the potential of RSOM to assess tumor angiogenesis in animals and visualize the microvasculature of human skin, *in vivo*, using only endogenous contrast [29, 30, 33-37]. For example, RSOM can image the internal and surrounding vasculature of a melanoma tumor in a mouse, *in vivo*, at depths up to 3 mm [30]. RSOM has also been applied to visualize skin morphology and vascular patterns in the dermis and sub-dermis of psoriasis patients, enabling computation of several biomarkers to differentiate between psoriasis and healthy skin [29, 31]. However, current RSOM systems can only scan a 4 × 2 mm^2^ area in 70 seconds due to the maximum repetition rate of the laser (500 MHz), which is determined by the maximum permissible exposure of human skin to laser light [29, 38]. This long scanning time makes RSOM susceptible to artefacts due to breathing or other motions. Motion correction algorithms reduce artefacts, but are insufficient to compensate for the large displacements that occur at the back and chest during breathing [39, 40]. This weakness is problematic for imaging nevi or melanoma, which are frequently found on a person’s upper torso, particularly the upper back.

We introduce here a fast RSOM (FRSOM) system based on a high-sensitivity ultrasound detector and a top coaxial illumination scheme, implemented by transmitting light using a single fiber through a central aperture of the high-sensitivity ultrasound transducer. These hardware innovations enable the use of a lower energy laser source at a high repetition rate, affording an almost four-fold decrease in scan-time, without sacrificing image quality or exceeding laser safety limits. FRSOM can scan a 4 × 2 mm^2^ field-of-view in only 15 seconds, which is short enough for a patient to hold his or her breath during imaging of the back and chest. We apply single-breath-hold FRSOM to visualize the microvasculature of human melanoma and surrounding skin tissue at unprecedented resolution-to-depth ratios. We demonstrate that biomarkers such as vascular density, complexity, and tortuosity can be extracted from single-breath-hold FRSOM images and used to distinguish groups of cutaneous malignant melanomas and benign nevi. This study introduces FRSOM vasculature imaging as a noninvasive method to provide complementary information to dermoscopy and increase the accuracy of melanoma detection.

## Results

### FRSOM system

The FRSOM system combines a custom-made spherically focused through-hole transducer with a coaxial illumination scheme (Fig. 1a, see also Methods), achieved by inserting a single fiber through the central aperture of the transducer (Supplementary Fig. 1). Compared to conventional RSOM, the coaxial illumination transmitted through a single fiber allows a 4-fold reduction in pulse energy and a concomitant increase in the laser repetition rate to 1.4 kHz (from 500 Hz), without decreasing the light fluence or exceeding laser safety limits (see Methods). In addition, an integrated miniature preamplifier was connected directly to the sensor circuits in the through-hole transducer, which minimized artifacts and doubled the FRSOM system’s achievable SNR compared to the transducer used in conventional RSOM (see Methods and Supplementary Fig. 1). This combination of a high-repetition laser with a more sensitive transducer allows FRSOM to scan an area of 4 × 2 mm^2^ in 15 seconds, with image quality comparable to that of conventional RSOM images recorded in 70 seconds. The full comparisons between the FRSOM and conventional RSOM were conducted by measuring the same skin tissue of a healthy volunteer and corresponding results are shown in Supplementary Fig. 1 and a volumetric FRSOM image is shown in Supplementary Video 1.

**Figure 1.**
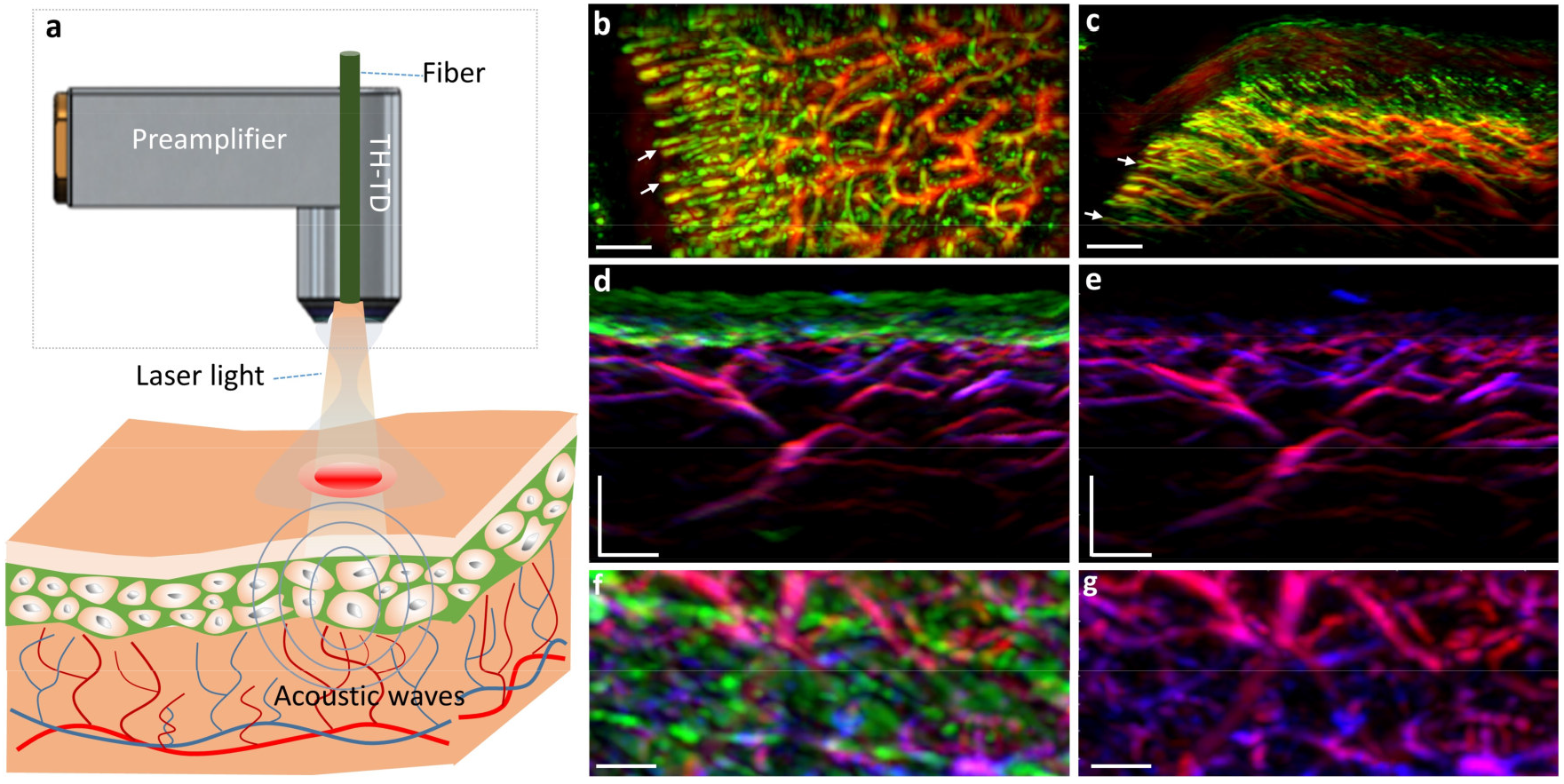
Schematic of the FRSOM system. a, Schematic of the FRSOM setup. The illumination scheme is implemented by transmitting light through the center aperture of the through-hole transducer (TH-TD) using a single fiber, which generates an illumination spot of 1.4 mm in diameter. b,c, MIP (maximum intensity projection) images in the cross-sectional and coronal direction of the microvasculature networks from the cuticle of a healthy volunteer; White arrows indicate the capillaries. d,f, MIP images in the cross-sectional and coronal direction of healthy skin structures recorded by multispectral FRSOM, where the reconstructed images were unmixed for melanin (green), deoxyhemoglobin (blue), and oxyhemoglobin (red). e,g, MIP images of deoxyhemoglobin (blue) and oxyhemoglobin (red) when the melanin channel was switched off of images (e and g); All scale bar: 500 µm.

To demonstrate the imaging performance of FRSOM, the microvasculature network of a volunteer’s cuticle was measured, and corresponding coronal and cross-sectional MIP (maximum intensity projection) images were shown in Fig. 1b-c. The capillaries (white arrows in Fig. 1b-c) in these images are clearly resolved with comparable image quality to that of conventional RSOM [35]. In addition, a multispectral FRSOM implementation was devised using a high repetition rate laser (1.4 kHz) with low pulse energy (see Methods). This multispectral FRSOM recorded a 4 × 2 mm^2^ section of healthy skin with four wavelengths (532, 555, 579, and 606 nm) in 60 seconds, compared to 13 minutes for a conventional multispectral RSOM setup [37]. The reconstructed images were unmixed to highlight melanin (green), deoxyhemoglobin (blue), and oxyhemoglobin (red, Fig. 1d-e), achieving comparable unmixing performance of our conventional spectral RSOM [37]. To visualize the location of melanin, the green channel (in which melanin absorbs) was switched off in the images of Fig. 1e and 1g, allowing the missing melanin features to be easily identified in the superficial skin layer compared with the images of Fig. 1d and 1f. Since the 60 second measurement time of this spectral FRSOM implementation exceeds a single-breath-hold, a 25 second dual-wavelength (515 nm and 532 nm) version (see Methods) was also tested and shown to discriminate the melanin and hemoglobin distributions of skin tissue (see Supplementary Fig. S2).

### Efficacy of motion suppression in single-breath-hold FRSOM imaging

Having established the imaging performance of FRSOM within a 15 second window, we sought to determine if a single-breath-hold by the patient during this time sufficed to suppress motion artefacts. To this end, we measured the healthy skin on the back of a female volunteer during both normal breathing and a 15 second single-breath-hold (Fig. 2). Fig. 2a-b show the raw optoacoustic signals of the FRSOM scans recorded with and without breathing motion, with the dashed lines indicating the skin surface. The recorded FRSOM data reveals the severe motion (maximum of 300 µm, beyond the capability of motion correction algorithms) caused by breathing (Fig. 2c), as well as the significant reduction in this motion (maximum of 70 µm) when the breath is held for 15 seconds (Fig. 2d, see Methods). Fig. 2e-f show the corresponding cross-sectional MIP images with and without breathing. Breathing motion causes extreme blurring of vascular structures (Fig. 2e), which cannot be corrected by any motion correction algorithms, whereas the volunteer simply holding her breath for 15 seconds enables the collection of high-resolution images of the skin vasculature (Fig. 2f).

**Figure 2.**
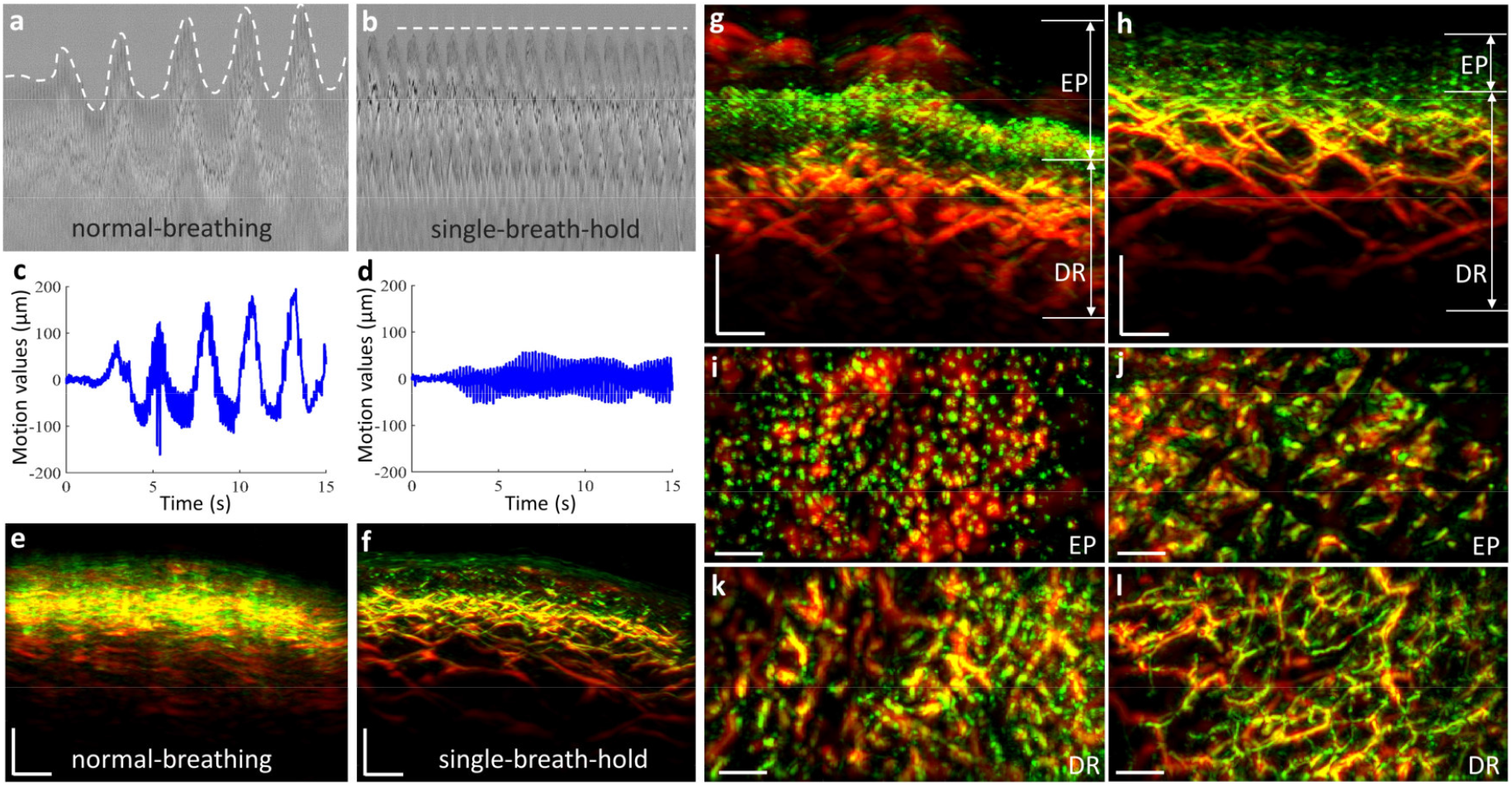
Single-breath-hold FRSOM imaging of healthy and psoriatic skin. a,b, Raw optoacoustic signals from FRSOM measurements of healthy skin on the back of a human volunteer, acquired during (a) normal breathing and (b) while the breath was held for 15 seconds (single-breath-hold). Dashed lines mark the positions of the skin surface in all A-line optoacoustic signals, which reflect the breathing motion. c,d, Corresponding motion graphs of (a) and (b), which show significantly different motion patterns between normal breathing and the held breath. e,f, Cross-sectional MIP images of healthy skin on the back of a human volunteer, acquired during (e) normal breathing and (f) while the breath was held for 15 seconds (single-breath-hold). A marked improvement in image quality is achieved when the breath is held during scanning compared to when normal breathing motions occur. g,h, Cross-sectional MIP images of psoriatic skin and adjacent healthy skin from the same volunteer’s back. i,j, Corresponding MIP images in the coronal direction of the epidermis (EP) layers of (g) and (h). k,l, MIP images in the coronal direction of the vascular networks in the dermis (DR) layer of (g) and (h). All scale bar: 500 µm.

The performance of FRSOM was assessed further by imaging a psoriasis lesion and the surrounding healthy skin (Fig. 2g-l) on the same patient’s back during 15 second single-breath-holds. The features of psoriatic skin at the human arm area were previously characterized using RSOM and provide a useful benchmark for the capabilities of FRSOM [29]. Fig. 2g reveals that FRSOM can resolve and quantify known features of psoriatic skin, including elongated and dilated capillary loops (visualized in green) extending through the rete ridges almost to the skin’s surface. Coronal FRSOM images close to the surface of the psoriatic skin (Fig. 2i) show the ends of the capillary loops, which appear markedly different from those in the corresponding image of healthy skin (Fig. 2j). In addition, the dermal vessels of the psoriatic skin (Fig. 2k) have larger diameters and appear denser than in the healthy skin (Fig. 2l). Overall, the previously documented epidermal thickening, capillary elongation, and increased dermal vascularization were clearly visualized by FRSOM during the 15 second breath-hold period.

### FRSOM vascular imaging of melanoma and surrounding skin tissue

We thus far demonstrated that FRSOM resolves morphological and vascular features of healthy and psoriatic skin on a patient’s back during a single-breath-hold, with performance similar to that of conventional RSOM when applied to measurements on a human arm where motion is minimal. To further elucidate the suitability of FRSOM for clinical applications, the system was used to visualize changes in skin microvasculature that had been altered by the progression of malignant melanoma. A melanoma lesion from a patient’s back was scanned during 15 second breath-holds in three regions (Fig. 3a, 1-3), each with a field view of 4 × 2 mm^2^ (red rectangle): the lesion base (scan 1), the lesion edge (scan 2), and the surrounding skin tissue (scan 3). A histological image from the lesion base (scan 1) is shown in Fig. 3b. Cross-sectional FRSOM images and the corresponding MIP images in the coronal direction of the epidermis (EP) and dermis (DR) layers are shown in Fig. 3c-k. Volumetric reconstructions of the three scans are presented in supplementary videos S2-S4.

**Figure 3.**
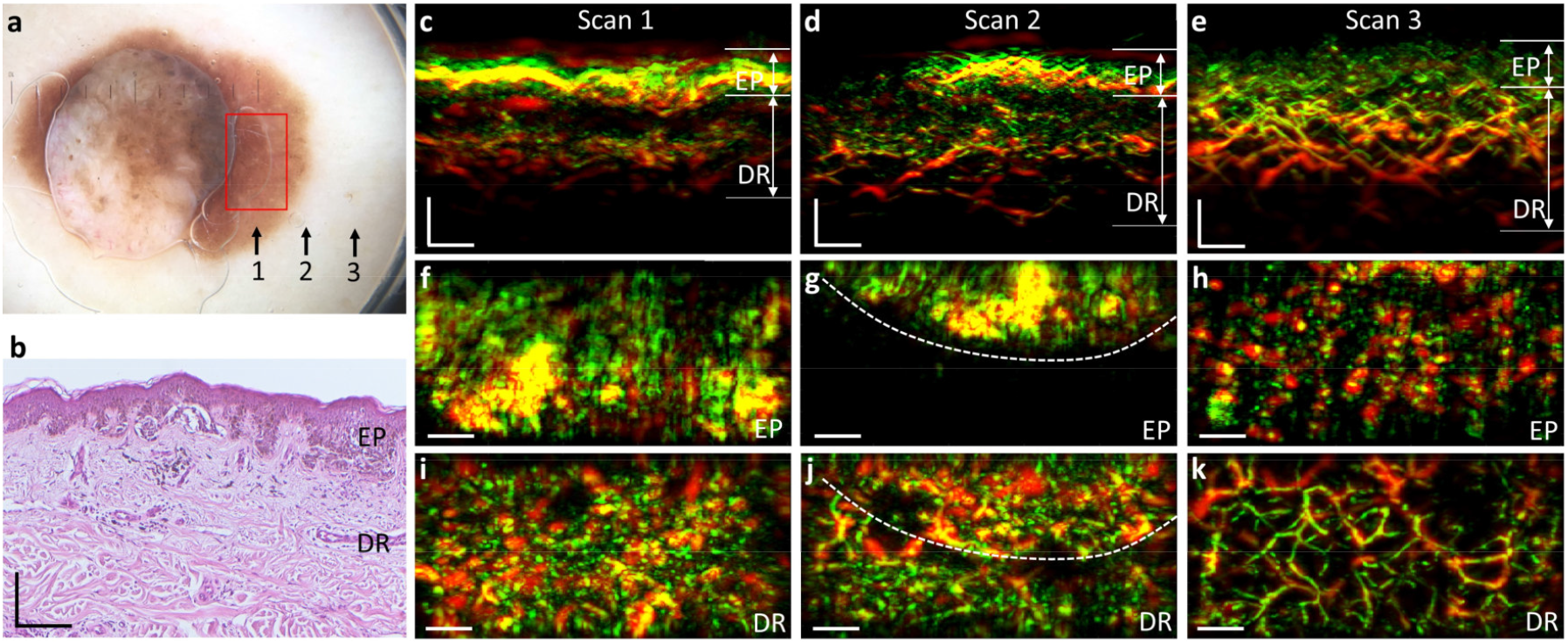
Vascular imaging of melanoma and the surrounding skin tissue. a, Photograph of a melanoma lesion; Black arrows and labels (1, 2, 3) indicate the three scanning positions, corresponding to the tumor base, the lesion edge and the surrounding skin tissue. The red rectangle depicts the field-of-view of 4 × 2 mm^2^. b, Histological image of the melanoma sample corresponding to the area (label 1) marked by the red rectangle in (a). c-e, MIP cross-sectional FRSOM images of the three scanned regions (labels 1, 2, 3). f-g, MIP images in the coronal direction corresponding to the epidermal (EP) layer of (c-e). i-k, coronal images corresponding to the dermal (DR) layer of Fig. 3c-e. The white dashed lines in (d) indicate the border between the pigmented lesion and the close surrounding skin tissue. Comparing the dermal vascular structures of (i-k), a dense dotted vascular pattern is clearly visualized in the tumor base area (i); irregular vasculature (j) is also resolved in the lesion edge area, while the vasculature networks become more regular in the surrounding skin tissue (k).

To illustrate how FRSOM resolves the vascular characteristics of the melanoma lesion, we compared the skin features in the epidermal and dermal layers at the three scan positions. The cross-section images (Fig. 3c-e) reveal the layered structure of the skin and the strong contrast of melanin in the EP layer, which clearly delineates it from the DR layer. Compared with the surrounding skin tissue, dermal vessels of the lesion appear irregular and disordered. Fig. 3f-h show coronal images corresponding to the EP layer in Fig. 3c-e. The coronal image of the lesion base (Fig. 3f) reveals the inhomogeneous melanin distribution within the melanoma. The boundary between the pigmented area and the adjacent tissue is clearly visible in the coronal image of the lesion’s edge (Fig. 3g, white dashed line). The healthy tissue surrounding the lesion is characterized by less contrast from melanin in the coronal MIP image (Fig. 3h) than in the lesion’s base and edge. Since angiogenesis is an important factor for tumour identification, we further investigated how the microvasculature features differed between the pigmented lesion areas and the surrounding skin tissue. Fig. 3i-k show coronal images corresponding to the DR layer of Fig. 3c-e. The dermal vasculature of the melanoma lesion and its edge exhibited a dense and dotted vessel pattern (Fig. 3i and 3j), characteristic of the disease [18, 41]. In contrast, entire vessels are discernible in the surrounding unaffected skin tissue (Fig. 3k). In the image of lesion’s edge (Fig. 3j), vascular patterns distinguish the pigmented lesion from the adjacent tissue (white dashed line), showing a transition of the vascular distribution from the highly dense and disordered pattern typical of melanoma to a regular vessel network. In addition, the microvasculature of the skin bordering the lesion (Fig. 3j) presented with irregular, dotted, and comma-like structures compared to the regular vascular network of skin farther from the lesion (Fig. 3k). Moreover, the overall vessel density decreases from the pigmented area of the lesion (Fig. 3i) to the lesion’s edge (Fig. 3j) to the tissue farther from the lesion (Fig. 3k).

### Quantification of FRSOM vasculature features between melanoma and dysplastic nevus

Quantifiable biomarkers can be extracted from RSOM’s high-resolution vascular images to provide objective measures of disease severity [29]. Here, we sought to evaluate this capability of FRSOM by quantifying and comparing pathophysiological features of benign dysplastic nevi and melanomas. For this, 10 dysplastic nevi and 10 melanomas were scanned on the upper torsos of ten patients by single-breath-hold FRSOM. Some lesions were measured at two different areas of the lesion’s edge, to afford 17 datasets for each of the nevus and melanoma groups. A photograph of a dysplastic nevus located on a patient’s back is shown in Fig. 4a; the red rectangle indicates the scanned region-of-interest at the edge of the lesion. A histological image and the corresponding cross-sectional FRSOM image are shown in Fig. 4b and 4c, respectively. A volumetric reconstruction of a nevus lesion is presented in supplementary videos S5. Fig. 4f shows an exemplary melanoma lesion and a red rectangle indicating the scanned region. The corresponding histology and cross-sectional FRSOM image are depicted in Fig. 4g and 4h, respectively. The dense pigmentation boundaries of the nevus and melanoma lesions appear in the coronal view MIP images of the EP layer (Fig. 4d and 4i, dashed lines). We observed dense and irregular vasculature, typical of melanoma [17, 20], in the dermis layer of the tissue immediately surrounding the melanoma (Fig. 4j). In comparison, the vasculature in the tissue surrounding the nevus appears comparable to healthy skin (Fig. 4e). We selected the vessels in the skin tissue just outside of the lesions (Fig. 4e,j, white dash lines) as regions-of-interest to compare the microvasculature between the nevus and melanoma, since the vasculature optoacoustic signals under the pigmented area may be attenuated by the melanin signals at the wavelength of 532 nm.

**Figure 4.**
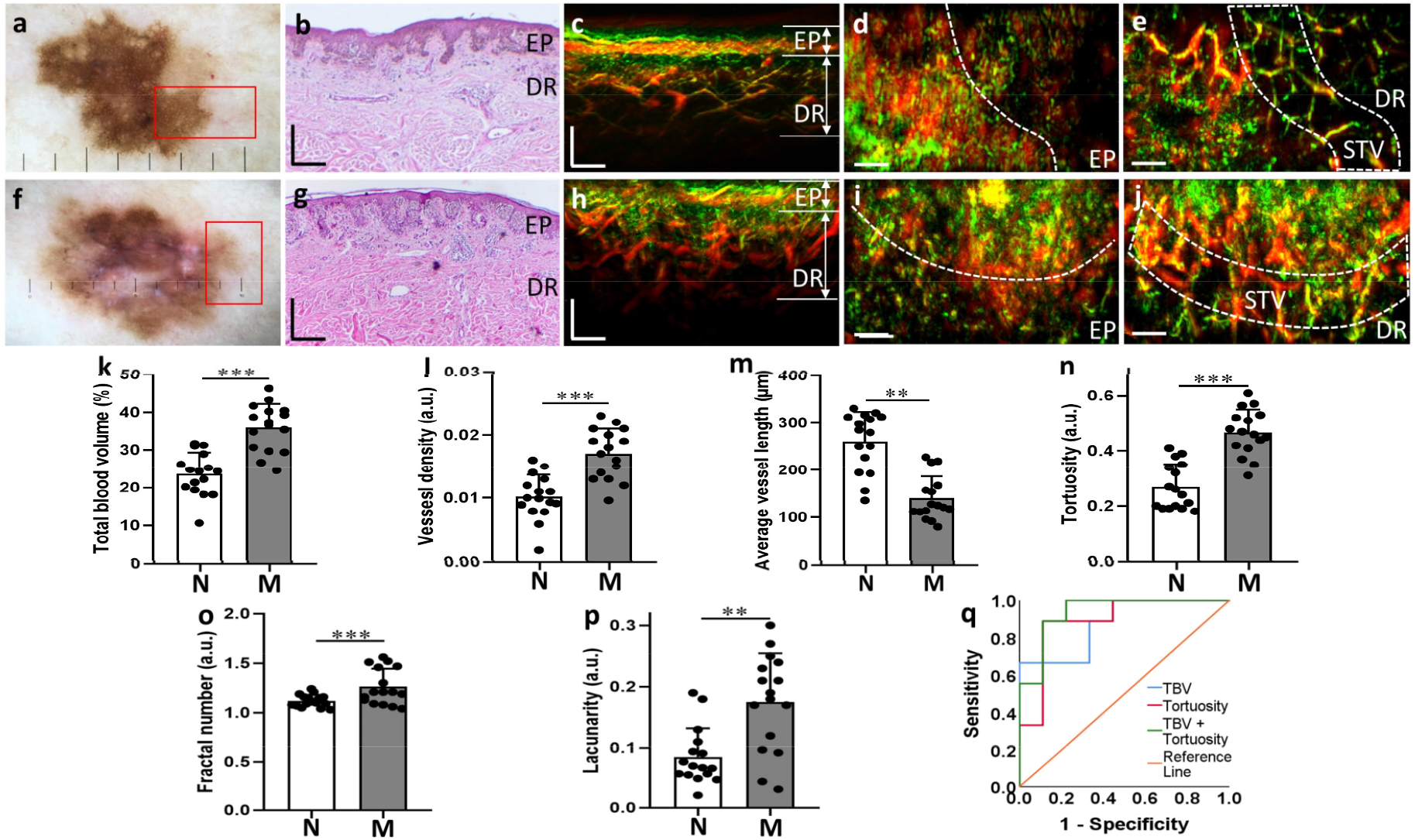
Vasculature feature quantifications between nevi and melanomas. a, Photograph of a dysplastic nevus from a patient back; the red rectangle indicates the scanning area. b, Histological image of the dysplastic nevus corresponding to the area marked by the red rectangle in (a). c, Cross-sectional MIP image measured at the edge area of the nevus marked by the red rectangle in (a). d,e, Corresponding MIP images in the coronal direction of the epidermis (EP) and dermis (DR) layers of (c). f, Photograph of a melanoma from a patient back; g, Histological image of the melanoma corresponding to the area marked by the red rectangle in (f). h, Cross-sectional MIP image measured at the edge area of the melanoma marked by the red rectangle in (e). i,j, Corresponding MIP images in the coronal direction of the EP and DR layers of (h); White dash lines in (d,e) indicate the separation boundaries between the pigmented lesion and surrounding skin tissue in the nevus and melanoma. White dash windows mark the vessel structure of the surrounding skin tissue, which is segmented by extending 500µm length from the separation dash lines towards the healthy skin. STV: surrounding skin tissue vessels. k-p, the computed vessel biomarkers: total blood volume (TBV), the vessel density, average vessel length, tortuosity, fractal number and lacunarity between the non-malignant nevi group (N, n=17) and melanoma group (M, n=17). q, ROC plots in the differentiation of melanoma using TBV, tortuosity, and the combination (TBV + tortuosity). **, and *** represent P < 0.01, and P < 0.001 respectively. All scale bar: 500 µm.

As a next step, we quantified vasculature features of the dysplastic nevi and melanomas to identify biomarkers for melanoma detection. The close surrounding tissue vessels (STV in Fig. 4e and 4j) in the dermis layers were segmented in a 500 µm distance extending out from the boundary of the pigmented lesion towards the healthy skin tissue. We then computed six biomarkers from these segmented vessels: (1) the total blood volume, (2) the vessel density, (3) the average vessel length, (4) the tortuosity, (5) the fractal number, and (6) the lacunarity (see Methods). The total blood volume and the vessel density metrics enable direct quantification of the amount of tumor-associated vascularity. The average vessel length and tortuosity indicated changes in the vascular geometry and morphology. The fractal number represents vascular spatial complexity associated with vascular changes and aberrant angiogenesis. Finally, the lacunarity characterizes tissue heterogeneity.

Fig. 4k-p show the results of the quantification and statistical comparisons of the six biomarkers. We observed significant differences of the six biomarkers characterized by the parametric tests (unpaired t test) for normally distributed data and nonparametric tests (Wilcoxon signed-rank). The dermal vasculature of the melanoma edge areas exhibited a significantly higher total blood volume and vessel density compared with the melanocytic nevus, showing much higher vascularity of melanoma compared to nevi. The average total blood volumes for the melanoma and nevi were 35.94% ± 6.22% vs. 23.63% ± 5.71%, respectively (Fig. 4k, *P* < 0.001) while the mean vessel densities were 0.017 ± 0.004 (a.u.) vs. 0.01 ± 0.003 (a.u.) (Fig.4l, *P* < 0.001). The average length of the segmented vessels was 260.39 ± 62.49 µm in the nevus group and 139.60 ± 46.63 µm in the melanoma group (Fig. 4m, *P* < 0.01), reflecting different spatial geometries of vessel patterns. The branched tree pattern of vasculature network was evident at the edge of the dysplastic nevi (Fig. 4e), while the vessel structure of the tissue surrounding the melanomas (Fig. 4j) was more tortuous and disorganized. These observations were confirmed by the tortuosity values of 0.27 ± 0.082 vs. 0.47 ± 0.083 arbitrary units (a.u.) for the nevus and melanoma groups, respectively (Fig. 4n, *P* < 0.001). The melanoma lesions exhibited a higher mean fractal number of 1.26 ± 0.18 a.u., compared with 1.12 ± 0.062 a.u. for nevus lesions (Fig. 4o, *P* < 0.001), measuring vascular spatial complexity associated with vascular changes and aberrant angiogenesis. The melanoma lesions exhibited a higher mean lacunarity value of 0.170 ± 0.083 a.u., compared with 0.088 ± 0.050 a.u. for nevus lesions (Fig. 3p, *P* < 0.01), indicating that the vasculature of melanoma is more inhomogeneous. In addition, we examined whether the combination of markers was useful in its ability to differentiate melanoma from nevus. Receiver operating characteristic (ROC) curves (Fig. 4q) were constructed based on the biomarkers of the TBV and tortuosity, revealing an area under the ROC curve (AUC) of 0.87 and 0.89 respectively, while the combinations of these two biomarkers achieved an area under the ROC curve of 0.93.

## Discussion

Skin microvasculature may have prognostic value in melanoma detection or determining its malignancy [18, 42], however, conventional imaging techniques lack the capabilities to fully exploit the microvasculature of melanoma lesions for these purposes. RSOM resolves human skin microvasculature morphology and quantifies biomarkers of skin diseases non-invasively and *in vivo* at greater depths and higher spatial resolutions than other optical methods. In this work, we constructed a fast RSOM (FRSOM) system capable of 4 times faster scanning than conventional RSOM, without a discernable loss of image quality or exceeding laser safety requirements, enabling single-breath-hold measurements and a significant reduction in motion artifacts. With the FRSOM, we for the first time comprehensively observed the microvascular morphology of melanoma on the human upper torso. Furthermore, our results demonstrate the ability of FRSOM to quantify the vascularity features in the adjacent tissue of melanoma, which had significantly increased density, complexity, and tortuosity compared to benign nevi.

Our previous implementation of RSOM scanned an area of 4 × 2 mm^2^ in approximately 70 seconds because safety limits capped the maximum laser repetition rate at 500 Hz for a wavelength of 532 nm. However, FRSOM employs a high-sensitivity ultrasound detector and a top coaxial illumination scheme to reduce the pulse energy of the illumination source by a factor of 4, affording a concomitant decrease in scan time to 15 seconds without exceeding laser safety limits. Therefore, FRSOM scans in a short enough time window for the patient to hold his or her breath, allowing the back and chest areas, which are severely impacted by breathing motion, to be imaged. As shown in Fig. 1 and supplementary figure S1, the FRSOM demonstrated identical image quality and resolved similar skin morphology compared to the conventional RSOM. Moreover, single-hold-breath FRSOM (Fig. 2) enabled quantification of vasculature features of healthy and psoriatic skin at back and chest areas with strong motions beyond the capability of conventional RSOM with standard motion correction algorithm [39]. Several studies reported irregularly distributed vessel patterns of melanoma measured by dynamic optical coherence tomography. However, the reliable evaluation of the vascular morphology in melanomas based on OCT is limited to the superficial dermal layer at a depth of up to 500 µm [17, 43]. FRSOM for the first time enables the microvasculature visualization of human melanoma and adjacent skin tissue on the upper torso at unprecedented resolution and depth up to 1.5 mm at the wavelength of 532 nm. The significantly irregular and disordered vasculature architecture of melanoma is clearly resolved compared to the surrounding skin tissue as shown in Fig. 3 and supplementary videos S2-S4.

We further evaluated the feasibility of FRSOM to detect melanoma by quantifying and comparing the vasculature biomarkers from 10 melanomas and 10 nevi. FRSOM images clearly revealed the adjacent skin tissue vasculature of melanoma to have more irregular vascular distribution compared to the nevi. Histological studies have revealed increased vascularity in melanomas when compared with nevi [18, 20, 22]. Likewise, two biomarkers (total blood volume and vessel density, as shown in Fig. 4k and 4l) computed from the *in vivo* FRSOM images showed significantly higher values in the tissue surrounding the melanomas than the nevi. In addition, previous OCT studies showed that melanomas display a more chaotic architectural organization of the superficial dermis layer in comparison to benign nevi [17, 44]. Similarly, four FRSOM biomarkers (the average vessel length, tortuosity, fractal number, and lacunarity) calculated for the deeper dermal vasculature quantified obvious morphological differences between the melanoma and benign nevus groups. The comparisons of the six FRSOM biomarkers provides an initial set of vasculature features that are characteristic of melanoma and benign nevi, corresponding well to descriptions from conventional histopathology and OCT studies [17, 44]. Therefore, FRSOM enables rapid, *in vivo*, noninvasive visualization of the microvascular architecture of melanoma lesions and adjacent skin tissue, which could increase the accuracy of melanoma detection and minimize the need for invasive biopsies. Furthermore, the increasing use of antiangiogenic drugs will stimulate demand for reliable, reproducible, and standardized means of assessing tumor angiogenesis response to these therapies [21]. FRSOM may provide a non-invasive alternative to repeated biopsies to assess the tumor vascularization of melanoma and monitor the response to antiangiogenic therapies [45].

The performance of multi-spectra RSOM is limited by unavailability of portable ultra-fast pulsed lasers with pulse widths of 1–3 ns at sufficient energy. Spectral RSOM imaging has been demonstrated in healthy skin, but at a lower resolution and speed due to employment of slow parametric oscillator lasers with pulse widths >10 ns. For example, a scan of 4 × 2 mm^2^ by our previous spectral RSOM implementation required approximately 13 minutes to record 4 wavelengths. In this work, the high sensitivity transducer with preamplifier and coaxial fiber illumination allows implementation of spectral FRSOM by using four-wavelength laser source (1.4 kHz) with low pulse energy (10-25 µJ), which is significantly faster than our previous implementation using slow parametric oscillator laser at a repetition rate of 100 Hz [37]. We show that a 60 second scan with FRSOM resolves the distributions of melanin, deoxyhemoglobin, and oxyhemoglobin in healthy skin (Fig. 1d-g). Because this time frame is still too long for the measurement with single-breath-hold, we implemented dual-wavelength (515 nm and 532 nm) FRSOM to discriminate the melanin and hemoglobin distributions of healthy skin, nevi, and melanomas (supplementary figure S2). Resolving melanin and hemoglobin can aid in identifying the boundary of a lesion for better selection of the possible excision margins (supplementary figure S2). The dual wavelength FRSOM took about 25 seconds to scan the area of 4 × 2 mm^2^, which enables single-breathing-hold spectra FRSOM measurements. Progress in ultra-sensitive ultrasound transducer and fast laser technology are essential to further decrease the acquisition time for single-breathing-hold spectral FRSOM.

The results of our study are limited by the small number of subjects. Benign nevi, in particular, have very diverse features and a larger population is necessary to validate diagnostic performance. A more comprehensive study of a larger number of patients is necessary to validate the computed biomarkers and evaluate how well FRSOM features can distinguish melanoma from benign nevi. In addition, we investigated the vascular features of the close surrounding skin tissue of the melanocytic lesions, as the penetration depth of FRSOM is limited to a depth of about 1.5 mm with a laser wavelength of 532 nm, which further decreases when imaging the pigmented areas due to strong melanin absorption (Fig. 3). Furthermore, it is difficult to separate the optoacoustic signals of vessels from the melanin signals. In order to better visualize and quantify the vasculature under the pigmented areas, an optical wavelength that has relative high hemoglobin absorption and low melanin absorption or spectra FRSOM can be employed to minimize the melanin signals and further improve the depth of melanoma vascular imaging.

In conclusion, we built a single-breath-hold FRSOM imaging system, which enables non-invasive visualization of the microvasculature of pigmented melanocytic lesions. The quantitative assessment of the vascular biomarkers exhibited significant differences of the vascular morphology between the nevi and melanoma groups. Although *in vivo* analysis and larger-scale studies are required to further validate the technique’s capability and feasibility, we believe that the results of our initial pilot study show great potential for FRSOM imaging as a preoperative screening tool, providing complementary information for melanoma detection as well as in the longitudinal monitoring of treatment options.

## Methods

### The FRSOM system

The FRSOM system (Fig. 1a) was implemented based on a custom-made spherically focused LiNbO_3_ through-hole transducer (TH-TD, Fig. 1b) and a coaxial illumination scheme achieved by inserting a single multi-mode fiber (Thorlabs, 200 µm in diameter) through the central aperture (350 µm in diameter) of the transducer. In our previous implementation of RSOM, the bundle illumination scheme (Supplementary Fig. 1a) achieved a homogeneous illumination pattern with diameter of 2.5 mm and restricted the repetition rate of laser source at the wavelength of 532 nm under 500 Hz because of the human-use safety limits described in the American National Standard for Safe Use of Lasers (ANSI Z136.1-2014, Laser Institute of America)[30]. However, the coaxial illumination through single fiber of FRSOM obtained a circle illumination pattern of 1.4 mm in diameter (Supplementary Fig. 1b) and enabled to reduce the pulse energy by a factor of 4 at a value of 18 μJ and correspondingly increased the laser repetition rate of 1.4 kHz without dropping the light fluence within the illuminated area. In addition, the novel TH-TD transducer was designed and implemented by directly mounting an integrated miniature preamplifier (30 dB amplification, ERA-8SM+, Mini-Circuits, Brooklyn, New York) on the ultrasound transducer before the signal was transmitted to the acquisition card (Gage, CSE161G2, 1 GS/s, US). In addition, we built an impedance matched transmission of the amplified signal that was less affected by the long transmission distance which gained two times of SNR by minimizing artifacts compared with the transducer without preamplifier used for the conventional RSOM system (Supplementary Fig. 1b-c). Our previous implementation of RSOM scanned over 4 mm × 2 mm (scan points, 266 × 135), taking 70 s. However, FRSOM images were reconstructed using optoacoustic signals collected over 4 mm × 2 mm (scan points: 201 x 101) with a total scanning time of 15 seconds, achieving about four times scanning speed acceleration. The imaging performance comparison between RSOM and FRSOM was characterized by measuring the same skin area at the forearm of a healthy volunteer sequentially as shown in Supplementary Fig. 1d-1i. We observed that the skin morphology (epidermis and dermis layers) and dermal vasculature were clearly resolved by both implementations with identical image quality. For the FRSOM vascular imaging, we use the single wavelength of a 532 nm laser (Bright Solutions, Italy) with temporal width of the pulse 0.9 ns and maximum repetition rate of 2 kHz. Besides, we further implemented multispectral FRSOM by using a custom-designed 4 wavelengths (532, 555, 579, and 606 nm) laser source with maximum repetition rate of 1.4 kHz and low pulse energy (10-25 µJ) at the fiber output. The multi-spectral FRSOM took about a total of 60 seconds to scan over 4 mm × 2 mm (scan points for each wavelength: 201 x 101), which exceeded a single-breath-hold. Further, dual-wavelength (515 nm and 532 nm) FRSOM was implemented to further reduce the scanning time of about 25 seconds for the area of 4 × 2 mm^2^ (scan points for each wavelength: 201 x 101). We have demonstrated that the dual-wavelength FRSOM can discriminate the melanin and hemoglobin distributions of healthy skin, nevus and melanoma as shown in the supplementary figure S2. The dual wavelength FRSOM could enable single-breathing-hold spectra FRSOM measurements for some individuals who could hold breath for 25 seconds.

### Motion correction and image reconstruction

Our previous developed motion correction method has been applied to correct motions of the recorded FRSOM data [39]. In Fig. 2a-d, the motion graphs between the normal-breathing and hold-breathing were computed by transforming the two dimensional motion matrix into one dimension. For the image reconstruction, the motion corrected FRSOM signals were divided into two frequency bands 10-40 MHz (low) and 40-120 MHz (high) for the 10-120 MHz bandwidth. Signals in the two different bands were independently reconstructed. The beam-forming method was used to reconstruct three-dimensional images [46]. The reconstruction algorithm was accelerated by parallel computing on a graphics processing unit and improved by incorporating the spatial sensitivity field of the detector as a weighting matrix. The reconstruction time of one bandwidth took about 2 minutes with voxel size of the reconstruction grid at 12 µm ×12 µm×3 µm. The two reconstructed images *R*_*low*_ and *R*_*high*_ corresponded to the low- and high frequency bands. A composite image was constructed by fusing *R*_*low*_ into the red channel and *R*_*high*_ into the green channel of a same RGB image. The detail process has been introduced in our previous work [29]. The FRSOM images can be rendered by taking the maximum intensity projections (MIPs) of the reconstructed images along the slow axis or the depth direction as shown in Fig. 1 and 2.

### Study population, histology, and general statistics

Twenty lesions (10 melanomas from 10 patients and 10 nevi from 9 patients) were imaged following approval from the Ethics Committee of Klinikum Rechts der Isar der Technischen Universität München, Munich, Germany. 10 lesions were located on back, chest or upper shoulder areas. The other 10 lesions were found on the arms and legs. In order to increase the sample size, 7 melanomas and 7 dysplastic nevi were measured two times at different edge areas of the lesion, generating 17 FRSOM datasets for the melanoma and nevus groups respectively. During the FRSOM scanning, patients were asked to hold breath for 15 seconds in order to reduce motions. Low quality datasets due to serious motion artefacts were excluded from the study.

All lesions were diagnosed by professional dermatologists and pathologists based on the histology image. CD31 immunostaining was also performed to evaluate vessel footprints. To assess the significance of the statistical differences for the metrics used to compare melanoma and nevus, we performed parametric tests (unpaired *t* test) for normally distributed data; otherwise, nonparametric tests (Wilcoxon signed-rank) were applied. Performing both tests ensured that the samples follow the required statistical distributions. The selection of the nevus and melanoma areas to be imaged was made by professional dermatologists, independently of the authors that processed the data. The diagnostic accuracy of independent marker or the combination of two markers was assessed with multivariate logistic regression and analyzed using ROC curves and by calculating the AUC.

### Lesion boundary and vessel segmentation, and biomarker calculation

With the 3D reconstructed FRSOM image, the skin surface was first flattened (Supplementary Fig. 3a). The corresponding MIP images in the coronal direction of the epidermis (EP) resolved the dense pigmentation distribution of the lesion. The epidermis layer in the MIP image of each data was automatically segmented by a graph theory and dynamic programming based approach [47] as marked by the dash lines (Supplementary Fig. 3b). Based on the segmented boundaries, we further separated the dermis vasculature (typically a volume of 4 mm × 2 mm × 2 mm was selected, although the dermis depth varied by patient) between the tumor base and the surrounding skin tissue (Supplementary Fig. 3c). The surrounding tissue vessels (STV) in the dermis layer was defined as 0.5 mm length extension from the boundary dash lines towards the healthy skin as shown in Supplementary Fig. 3d. To quantify the features of STV, we further segmented the vessel boundaries (Supplementary Fig. 3e), the skeleton and branching points (Supplementary Fig. 3e) based on the AngioQuant method [48].

The total blood volume (TBV) is determined as the ratio *TBV=N/V*, where *N* is the number of nonzero voxels from the STV image volume and *V* is the total segmented volume. The vessel density is calculated from the two-dimensional segmented STV image (Supplementary Fig. 2e) as the ratio between the total nonzero pixels of STV and the whole segmented area. The average vessel length is calculated as the ratio between the sum lengths of all vessel skeletons and the total number of branching points (indicated as the vessel number). The tortuosity of the curved vasculature is computed based on the distance metric (DM) defined as the ratio between the actual path length (AL) of a vessel and the straight-line distance (SL) between two end points of this blood vessel: 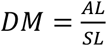 [49]. The fractal dimension number (FN) describes the complexity of an irregular object [50]: 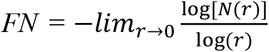, where *N(r)* represents the number of cubes required to cover an object when the cube side length is *r*. Generally, the higher the complexity of an object, the higher the fractal dimension number. In addition, the lacunarity (L) can be used to characterize the “lumpiness” of the fractal data, providing metainformation about the computed FN values [50], defined as: 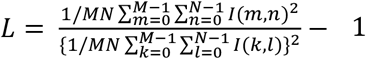, where *M* and *N* are the size of the FN processed image *I*. The higher the lacunarity, the more inhomogeneous the examined fractal areas and vice versa [50].

## Data Availability

The data that support the findings of this study are available from the corresponding author upon reasonable request.

## Acknowledgements

This project has received funding from the European Union’s Horizon 2020 research and innovation programme under grant agreement No 687866 (INNODERM) and under grant agreement No 871763 (WINTHER). We thank Dr. Robert J. Wilson for advice and editing and the staff at Department of Dermatology and Allergy at TUM for assisting with the patient studies.

## Author Contributions

H.H. developed the imaging system, designed and performed the experiments, processed the data, led the research, provided conceptual input and wrote the paper. C.S. recruited the patients, performed the patient experiments and provided conceptual input. M.S. developed the imaging system. B.H. recruited the patients and helped with the patient experiments. A.B. helped to perform the experiments. A.G. performed the histology experiments. U.D. provided conceptual input. J.A. provided conceptual input, helped to develop the imaging system, designed the experiments, performed the experiments, contributed to writing the paper and led the research. V.N. provided conceptual input, designed the experiments, supervised and led the research, and wrote the paper.

## Supplementary Information

**Figure S1.**
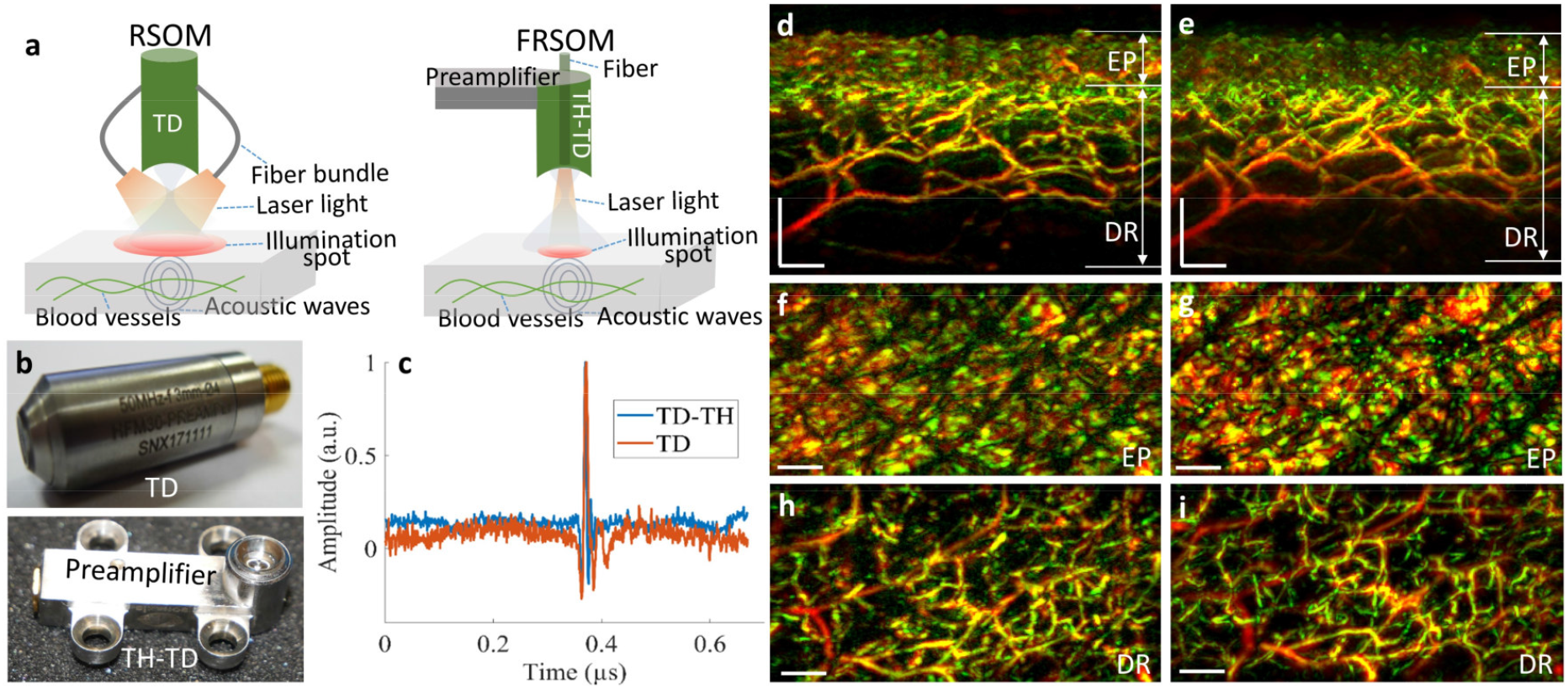
Comparison of conventional RSOM and FRSOM. a, schematic of the conventional RSOM and FRSOM. The conventional RSOM applied fiber bundle-based illumination with 2.5 mm diameter while the FRSOM employed the single fiber illumination with a 1.4 mm diameter. b, the normal transducer (TD) used in the conventional RSOM and the through-hole transducer (TH-TD) with preamplifier for the FRSOM. c, Signal quality comparisons between the TD and the TH-TD with preamplifier when measuring same object with external illumination. The TD-TH gains two times of SNR compared to the TD. d-e, Quality comparisons of the cross-sectional images between the conventional RSOM (d) and FRSOM (e) when measuring the same skin area at arm from a healthy volunteer. f-g and h-i are corresponding MIP images in the coronal directions from the epidermal (EP) and dermal (DR) layers respectively. We observed highly comparable image quality between the conventional RSOM and FRSOM.

**Figure S2.**
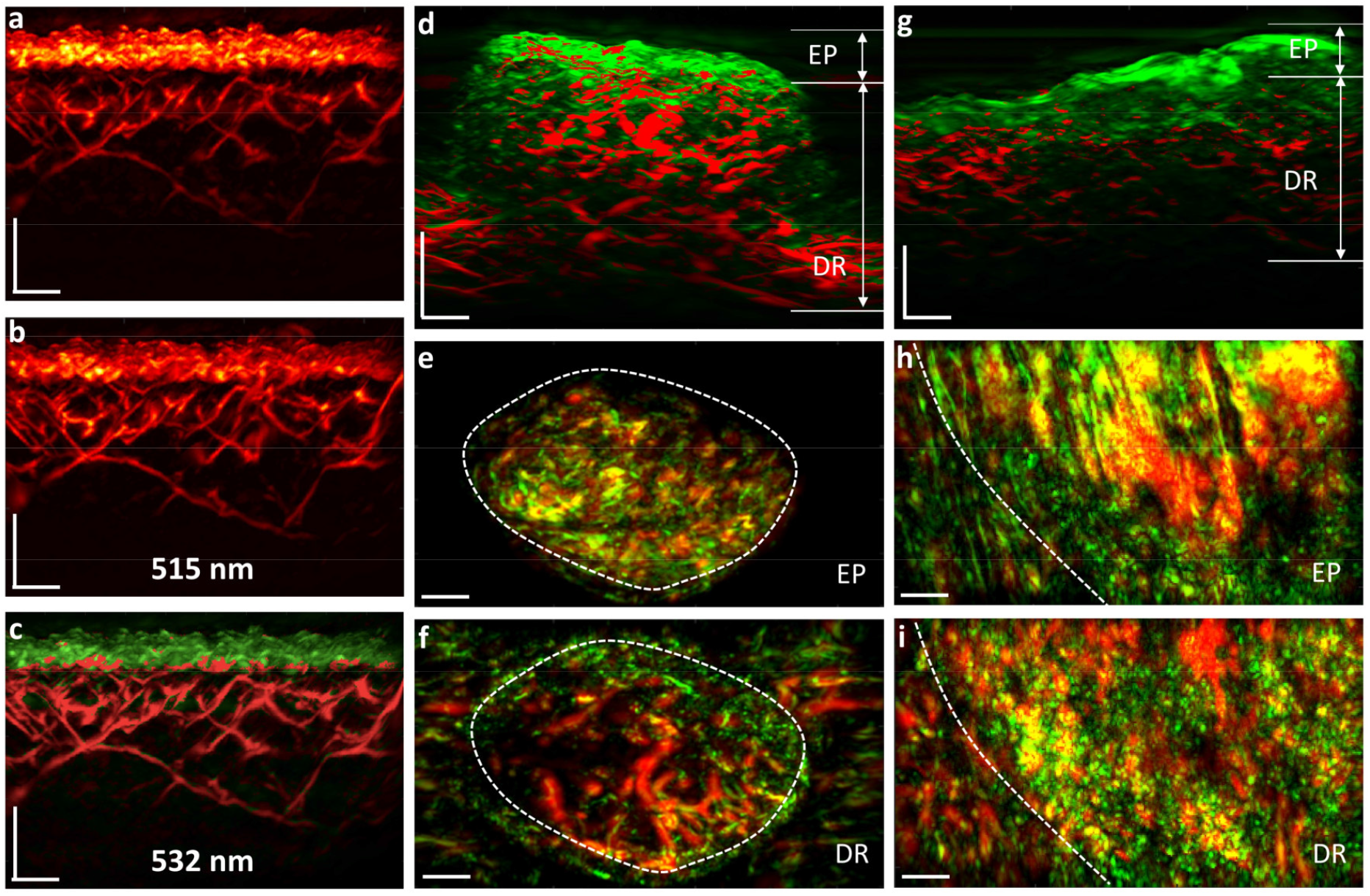
The dual-wavelength FRSOM imaging. a, MIP image in the cross-sectional direction of healthy skin recorded by the dual-wavelength FRSOM, where the reconstructed images were unmixed for melanin (green) and hemoglobin (red). b,c, MIP images in the cross-sectional direction recorded by FRSOM at a wavelength of 515 nm and 532 nm respectively. d, MIP image in the cross-sectional direction of a pigmented nevus recorded by the dual-wavelength FRSOM, where the reconstructed images were unmixed for melanin (green) and hemoglobin (red). e,f, MIP spectral FRSOM images in the coronal direction of the nevus at the epidermis (EP) and dermis (DR) skin layers. The white dashed lines indicate the pigmented lesion boundary. g, MIP image in the cross-sectional direction of a melanoma lesion recorded by dual-wavelength FRSOM, where the reconstructed images were unmixed for melanin (green) and hemoglobin (red). h,i, MIP spectral FRSOM image in the coronal direction of the melanoma at the epidermis (EP) and dermis (DR) skin layers. The white dash lines indicate the pigmented lesion boundary. Dual-wavelength FRSOM successfully differentiates melanin and hemoglobin structures in the healthy skin, nevus, and melanoma.

**Figure S3.**
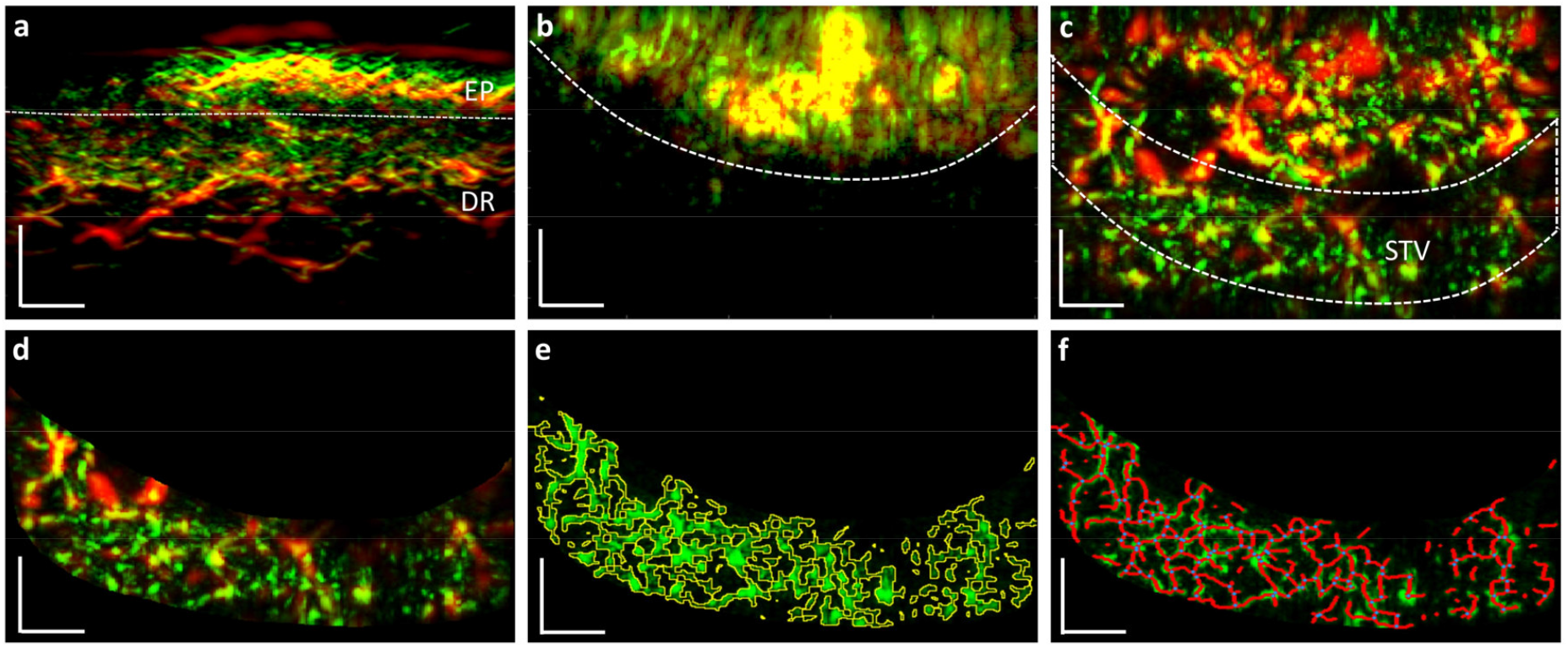
The FRSOM image and vessel segmentation procedures. a, cross-sectional FRSOM image of a melanocytic lesion, where the epidermal (EP) and dermal (DR) layers are delineated by the white dashed line. b, MIP image in the coronal direction corresponding to the epidermal layer of (a), where the dashed line indicates the boundaries separating the pigmented area of the lesion and the surrounding skin tissue based on the melanin contrast. c, MIP image of dermal vasculature corresponding to the dermal layer of (a), where the surrounding tissue vessel (STV), segmented from the boundary line to 500 μm thickness towards the healthy skin as indicated by the white dash box. d, The segmented image of the STV. e, the segmented vessel boundaries and the corresponding vessel centerlines (f).

